# AI-powered Coronary Artery Calcium Scans (AI-CAC) Enables Prediction of Heart Failure and Outperforms NT-proBNP: The Multi-Ethnic Study of Atherosclerosis

**DOI:** 10.1101/2024.01.04.23300650

**Authors:** Morteza Naghavi, Anthony Reeves, Matthew Budoff, Dong Li, Kyle Atlas, Chenyu Zhang, Thomas Atlas, Claudia Henschke, Christopher Defilippi, Daniel Levy, David Yankelevitz

**Author notes:** **Address for correspondence:** Morteza Naghavi, 2450 Holcombe, Houston, TX, 77021.

## Abstract

**Introduction:** Coronary artery calcium (CAC) scans contain useful information beyond the Agatston CAC score that is not currently reported. We recently reported that artificial intelligence (AI)-enabled cardiac chambers volumetry in CAC scans (AI-CAC) predicted incident atrial fibrillation in the Multi-Ethnic Study of Atherosclerosis (MESA). In this study, we investigated the performance of AI-CAC for prediction of incident heart failure (HF) and compared it with 10 known clinical risk factors, NT-proBNP, and the Agatston CAC score.

**Methods:** We applied AI-CAC to 5750 CAC scans of asymptomatic individuals (ages 45-84, 52% women, White 40%, Black 26%, Hispanic 22% Chinese 12%) free of known cardiovascular disease at the MESA baseline examination (2000-2002). We then used the 15-year outcomes data and compared the C-statistic of AI-CAC with NT-proBNP and the Agatston score for predicting incident HF versus 10 known clinical risk factors (age, gender, body surface area, diabetes, current smoking, hypertension medication, systolic and diastolic blood pressure, LDL, HDL, total cholesterol, and hs-CRP).

**Results:** Over 15 years of follow-up, 256 HF events accrued. The ROC area under the curve for predicting HF with AI-CAC (0.826) was significantly higher than NT-proBNP (0.742) and Agatston score (0.712) (p<.0001), and comparable to clinical risk factors (0.818, p=0.4141). AI-CAC category-free NRI significantly improved on clinical risk factors (0.43), NT-proBNP (0.68), and Agatston score (0.71) for HF prediction at 15 years (p<0.0001).

**Conclusion:** AI-CAC significantly outperformed NT-proBNP and the Agatston CAC score and significantly improved category-free NRI of clinical risk factors for incident HF prediction.

## Introduction

Coronary artery calcium (CAC) scoring is a strong predictor of coronary heart disease (CHD) events in asymptomatic individuals^1^. However, it is a weak predictor of heart failure (HF)^2^. HF is a significant healthcare burden due to large and rising numbers of HF hospitalizations and rehospitalizations^3^. Approximately 2% of the total US healthcare budget is spent on HF, and half of that is attributable to late diagnosis leading to inpatient admissions^4^. Numerous studies have highlighted the importance of early detection of asymptomatic high-risk patients to prevent progression to overt clinical HF. These high-risk patients include those with asymptomatic left ventricular dysfunction (ALVD), left ventricular hypertrophy (LVH), cardiomegaly, and enlarged cardiac chambers that if not detected and treated will lead to HF. They can be detected with imaging modalities such as echocardiography, cardiac computed tomography, and cardiac magnetic resonance imaging. Similarly, measurement of B-type natriuretic peptide (BNP) or the biologically inactive amino-terminal of BNP (NT-proBNP) that are circulating biomarkers of cardiac volume overload is associated with subclinical HF^5,6,7,8^. However, screening the asymptomatic population for detection of subclinical HF by any diagnostic means is not part of current guidelines due to cost and feasibility limitations^9^.

CAC scans contain insightful information beyond a CAC score that is not currently reported^10^. We recently reported that artificial intelligence (AI)-enabled volumetry of left atrium in CAC scans enabled prediction of incident atrial fibrillation in the Multi-Ethnic Study of Atherosclerosis (MESA) occurring within one year^11^. MESA is comprised of 6814 asymptomatic individuals free of known cardiovascular disease at baseline who were recruited for prospective evaluation in 2000-2002. In this study, we investigated the predictive value of AI-enabled cardiac chambers volumetry in CAC scans (AI-CAC) for prediction of incident symptomatic heart failure (HF). We further compared the predictive performance of AI-CAC with the best clinical model comprising 10 known risk factors (age, gender, body surface area, diabetes, current smoking, hypertension medication, systolic and diastolic blood pressure, LDL, HDL, total cholesterol, and hs-CRP) plus the addition of NT-proBNP or the Agatston CAC score.

## Methods

### Study population

The Multi-Ethnic Study of Atherosclerosis (MESA) is a prospective cohort study that began in July 2000 to investigate the prevalence, correlates, and progression of subclinical cardiovascular disease (CVD) in individuals without known CVD at baseline. The cohort included 6814 women and men aged 45–84 years old recruited from 6 US communities. Further detail has been published elsewhere^12^.

For our study, participants who did not consent to use of their data by commercial entities, cases with missing slices in CAC scans, and cases with missing NT-proBNP values were excluded from the analyses.

### Outcomes

Participants were contacted by telephone every 9-12 months during follow-up and asked to report all new cardiovascular diagnoses. International Classification of Disease (ICD) codes were obtained for hospitalizations. Reviewers classified HF as definite, probable, or absent. Definite or probable HF required heart failure symptoms, such as shortness of breath or edema, as asymptomatic disease is not a MESA endpoint. In addition to symptoms, probable HF required HF diagnosed by a physician and patient receiving medical treatment for HF. Definite HF required one or more other criteria, such as pulmonary edema/congestion by chest X-ray; dilated ventricle or poor LV function by echocardiography or ventriculography; or evidence of left ventricular diastolic dysfunction. The criteria for possible HF are a physician diagnosis of HF and receiving medical treatment for HF.

### The AI-CAC

The AI-enabled automated cardiac chambers volumetry tool in AI-CAC is called AutoChamber^TM^ (HeartLung.AI, Houston, TX), a deep learning model that used TotalSegmentator^13^ as the base input and was further developed to segment not only each of the four cardiac chambers; left atrium (LA), left ventricle (LV), right atrium (RA), and right ventricle (RV) but also ascending aorta, aortic root and valves, pulmonary arteries, and several other components which are not presented here. In this manuscript, the AI-estimated LA, LV, RA, RV, and LV wall volumes are referred to as AI-CAC or AI cardiac chambers volumetry, interchangeably.

Figure 1 shows the segmentations of cardiac chambers in color. The base architecture of the TotalSegmentator model was trained on 1139 cases with 447 cases of coronary CT angiography (CCTA) using nnU-Net, a self-configuring method for deep learning-based biomedical image segmentation^14^. The initial input training data were paired non-contrast and contrast-enhanced ECG-gated cardiac CT scans of the same individuals with 1.5 mm slice thickness. Because the images were taken from the same patients in the same session, registration was accomplished with good alignment. Following this transfer of segmentations, a nnU-Net deep learning tool was used for training the model.

**Figure 1.**
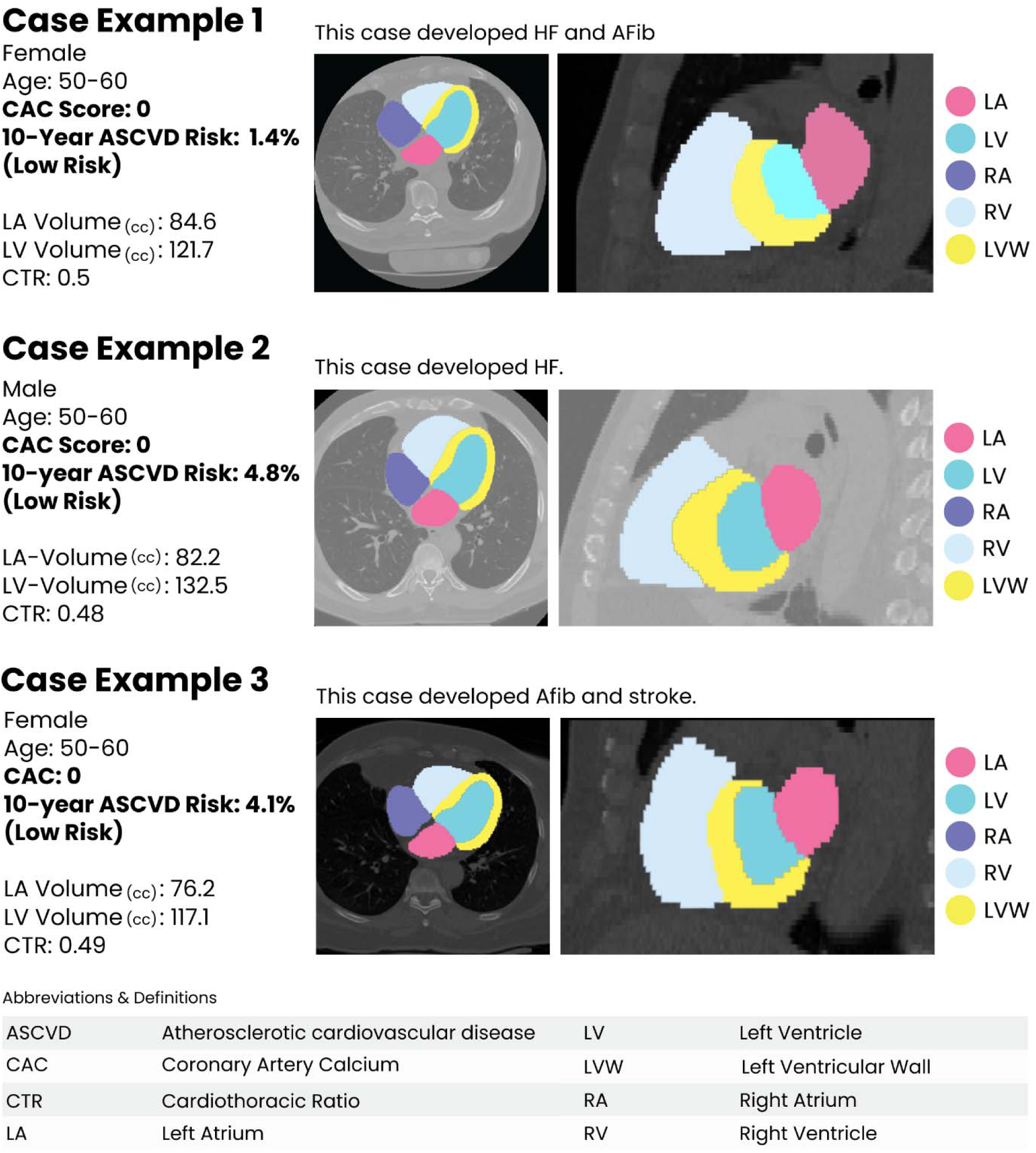
Examples of AI-CAC detection of high-risk individuals with enlarged left ventricle (LV) in coronary artery calcium (CAC) scans with calcium score of zero. The above images are created by AI-CAC from standard CAC scans. *Left atrium (LA), left ventricle (LV), right atrium (RA), right ventricle (RV), left ventricular wall (LVW), atrial fibrillation (Afib), cardiothoracic ratio (CTR)

Additionally, iterative training was implemented whereby human supervisors corrected errors made by the model, and the corrected data were used to further train the model, leading to improved accuracy. AI-CAC was run on 6043 non-contrast CAC scans that consented to commercial data usage out of the 6814 scans available in MESA exam 1. Expert rules built in the AI-CAC excluded 125 cases due to missing slices in image reconstruction created by some of the electron beam CT scanners used in MESA baseline.

### NT-proBNP

NT-proBNP data were obtained from MESA core laboratory for MESA exam 1 participants. A detailed study design for MESA has been published elsewhere^12^. Details on BNP assays used in MESA have been reported^15^. N-terminal proBNP is more reproducible than BNP at the lower end of the distribution range, and more stable at room temperature. However, both BNP and N-terminal proBNP are clinically available. Intra-assay and inter-assay coefficients of variation at various concentrations of NT-proBNP have been previously reported^16,17^. The analytical measurement range for NT-proBNP in exam 1 was 4.9–11699 pg/ml. The lower limit of detection for the NT-proBNP assay is 5 pg/mL, thus cases above 0 and below 4.99 were treated as 4.99 pg/mL. Numerous studies have established NT-proBNP as a predictor of incident HF, with a relatively high negative predictive value (94%–98%), but a significantly lower positive predictive value (44%–57% for nonacute heart failure, 66%–67% for acute heart failure)^18,19,20^.

### Statistical Analysis

We used SAS (SAS Institute Inc., Cary, NC) and Stata (StatCorp LLC, College Station, TX) software for our statistical analyses. All values are reported as means ± SD except for NT-proBNP which did not show normal distribution and is presented in median and interquartile range (IQR). All tests of significance were two tailed, and significance was defined at the p<0.05 level. The ROC (receiver operator curve) AUC (area under the curve) was calculated using Cox Proportional Hazards Regression. NT-proBNP values were right skewed and natural log converted to obtain a normal distribution for analyses. Category-free (continuous) net reclassification index (NRI) was calculated using the sum of the differences between the proportions of upward reclassifications and downward reclassifications for HF events and HF non-events, respectively. NRI was developed as a statistical measure to evaluate the improvement in risk prediction models when additional variables are incorporated into a base model^21^.

In our study, the base models are 10 known clinical risk factors (defined as age, gender, body surface area, diabetes,current smoking, hypertension medication, systolic and diastolic blood pressure, LDL, HDL, total cholesterol, and hs-CRP), NT-proBNP, and Agatston CAC score. The AI enabled cardiac chambers volumetry (AI-CAC) integrates all cardiac chambers (LA, LV, RA, RV, LV Mass) within the same model. Whereas AI-CAC LV specifically refers to AI-CAC of the left ventricle. We have analyzed data for HF prediction at 5, 10, and 15 years follow up.

## Results

For our study, we removed data from 771 MESA participants who did not consent to use of their data by commercial entities leaving 6043 participants at baseline. Among the remaining participants, after removing cases with missing data including 125 cases with missing slices in CAC scans and 168 cases with missing NT-proBNP values, the total number of cases available for analysis was 5750.

28.9% of participants were aged 45-54 years, 27.6% aged 55-64 years, 29.4% aged 65-74 years, and 14.1% aged 75-84 years. 53.3% were women, 38% were White, 28% Black, 22% Hispanic, and 12% Chinese. Table 1 shows the baseline characteristics of MESA participants who were diagnosed with incident HF versus those who were not over the period of 15 years follow up. At 5-, 10-, and 15-years follow-up 92, 188, and 261 HF cases were identified respectively. In univariate comparisons, incident HF cases were older, more likely male, and more likely White. The incident HF cases had higher cardiac chamber volumes for LA, LV, RA, RV, LV Wall, and NT-proBNP levels versus those without incident HF (all comparisons p< 0.001) (Table 1).

**Table 1:**
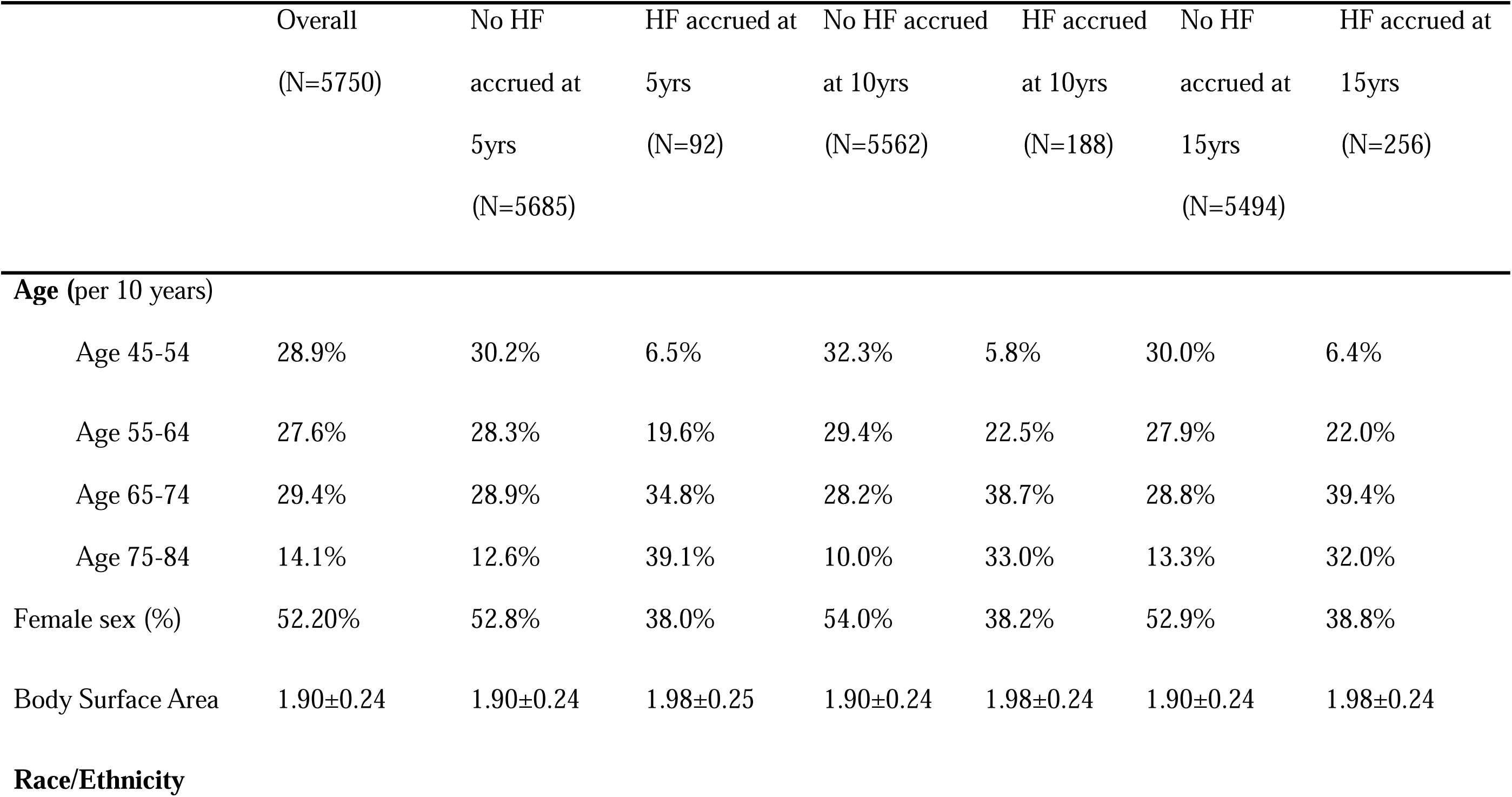
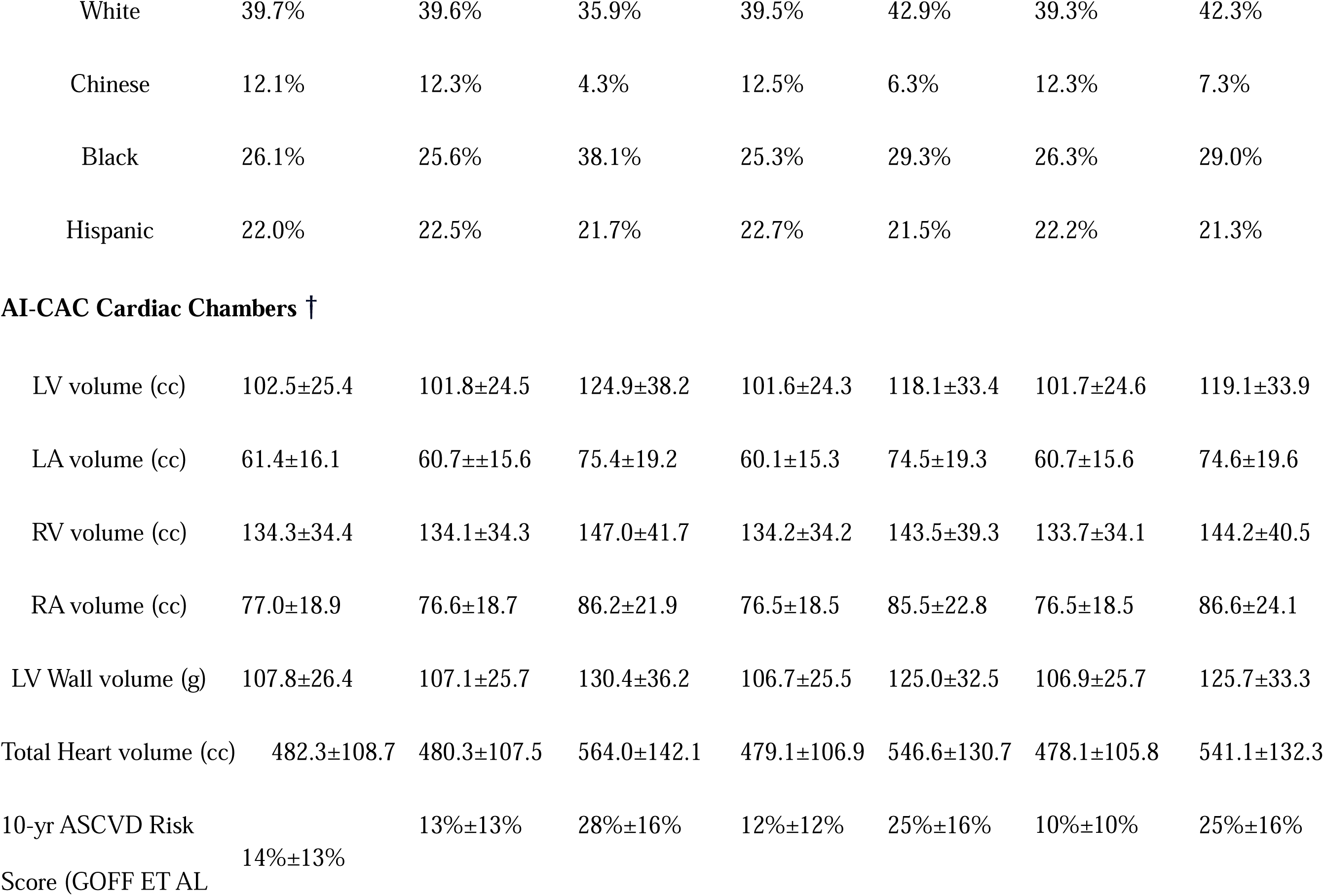
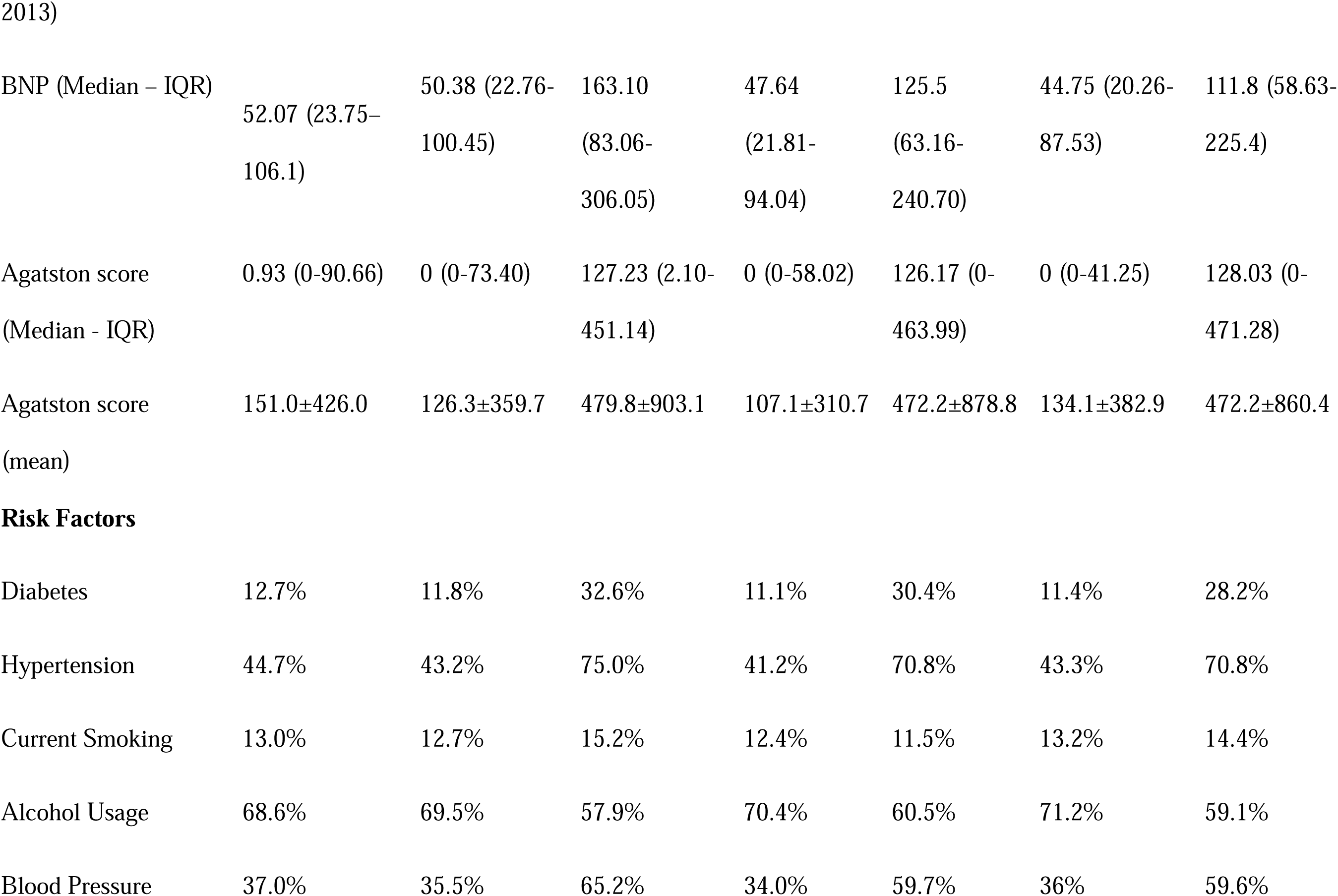
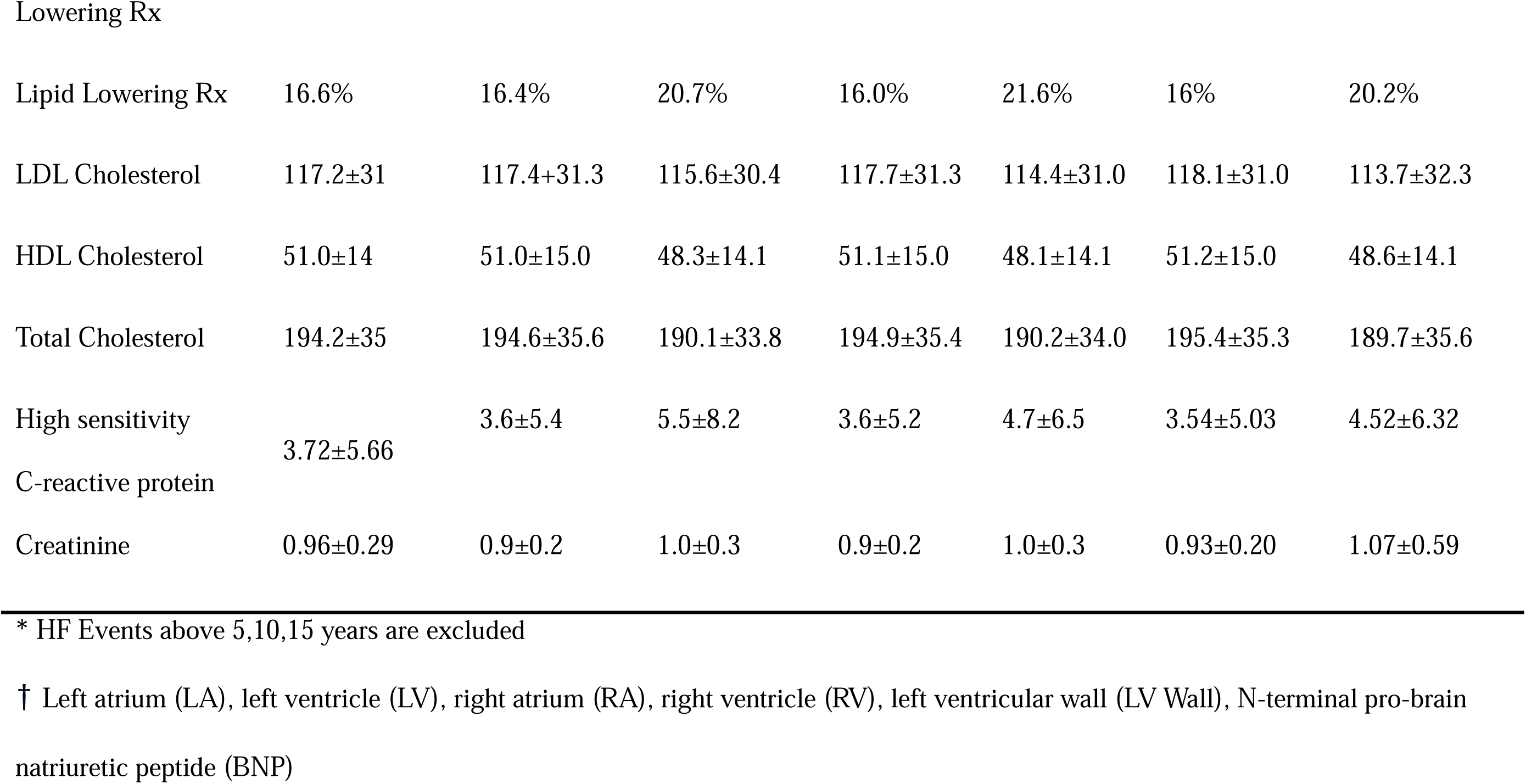
Baseline characteristics of the Multi-Ethnic Study of Atherosclerosis (MESA) participants including cases with and without heart failure (HF) at 5-,10-, and 15-years.

The AUC for AI-CAC adjusted by age, gender, and BSA was comparable to clinical risk factors model at 5-, 10-, and 15 years (0.838 vs. 0.824, p=0.58) (0.824 vs. 0.803, p=0.09) (0.826 vs. 0.818, p=0.41), respectively. AUC for AI-CAC was comparable to NT-proBNP for 5 years (0.838 vs. 0.794, p=0.21) and significantly higher than AUC for NT-proBNP for 10- and 15-year prediction of HF (0.824 vs. 0.745, p<.0001 and 0.826 vs. 0.741, p<.0001, respectively). AUC for AI-CAC versus the Agatston CAC score was 0.838 vs. 0.776 for 5 years (p<0001); 0.824 vs.

0.711 for 10 years (p<.0001); and 0.826 vs. 0.712 for 15 years (p<.0001). (Table 2).

**Table 2.** Comparing Area under Curve (AUC) for Heart Failure (HF) Prediction between Artificial Intelligence-enabled cardiac chamber volumetry (AI-CAC), Agatston CAC Score, NT-proBNP, and Clinical Risk Factors over 5, 10, and 15 Years in the Multi-Ethnic Study of Atherosclerosis (MESA)

|  | 5-year follow-up | 10-year follow-up | 15-year follow-up |
| --- | --- | --- | --- |
| HF Events accrued over follow-up | 92 | 188 | 256 |

| **AUC** | **AUC** | **P value** | **AUC** | **P value** | **AUC** | **P value** |
| --- | --- | --- | --- | --- | --- | --- |
| AI-CAC (AI-enabled All Cardiac Chambers Volumetry)* | 0.838 | - | 0.824 | - | 0.826 | - |
| Agatston CAC Score | 0.688 | 0.0004 | 0.710 | <.0001 | 0.712 | <.0001 |
| NT-proBNP | 0.795 | 0.2640 | 0.745 | <.0001 | 0.741 | <.0001 |
| Clinical Model† | 0.8242 | 0.5809 | 0.803 | 0.0857 | 0.818 | 0.4141 |
| AI-CAC LV Volumetry* | 0.818 | 0.3629 | 0.793 | 0.0231 | 0.804 | 0.0434 |

\*Adjusted by age, gender, body surface area (BSA)
† 10 known risk factors: Age, gender, BSA, diabetes, current smoking, hypertension medication, systolic blood pressure, diastolic blood pressure, high sensitivity C-reactive protein, LDL cholesterol, HDL cholesterol, total cholesterol

The AUC for AI-estimated LV volume alone (adjusted by age, gender, BSA) was comparable to NT-proBNP for 5 years (0.818 vs. 0.794, p=0.42) and significantly higher than AUC for BNP for 10- and 15-year prediction of HF (0.793 vs. 0.745, p=0.017 and 0.804 vs. 0.741, p<.0001, respectively) (Figure 2a-b). Despite Agatston CAC score not being relevant for HF prediction, the add-on to CAC scans makes the comparison insightful. AUC for AI-estimated LV volume versus CAC was 0.827 vs. 0.776 for 5 years (p=0.001); 0.789 vs. 0.711 for 10 years (p<.0001); and 0.819 vs. 0.712 for 15 years (p<.0001) (Table 2). The continuous NRI for 5-, 10-, and 15-years prediction of HF when AI-CAC was added to the Agatston CAC score as the only predictor in the base model was highly significant (0.81, 0.67, 0.71, respectively p<0.0001). The NRI for AI-CAC over 5, 10, and 15 years when added to base model NT-proBNP was significant (0.68, 0.71, 0.68, respectively, p for all < 0.0001). The NRI for AI-CAC over 5,10, and 15 years when added to the clinical risk factors model was 0.45, 0.50, 0.43, respectively, p for all <.0001. (Table 3).

**Figure 2a.**
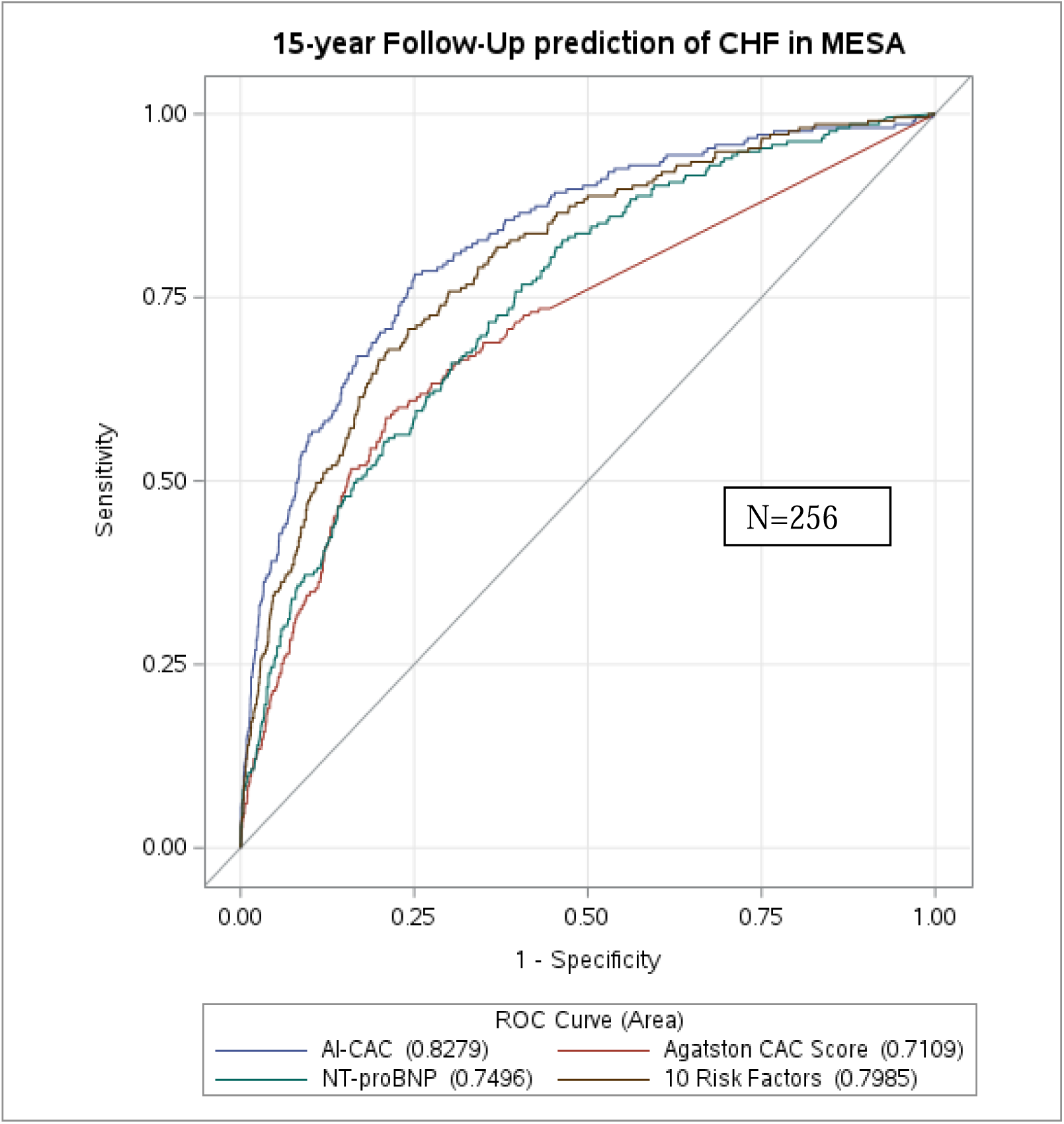
AI-CAC vs Agatston CAC score p<.0001; AI-CAC vs NT-proBNP: p<.0001; AI-CAC vs. 10 Risk Factors (p=0.4141) Clinical model (10 known risk factors): Age, gender, BSA, diabetes, current smoking, hypertension medication, systolic blood pressure, diastolic blood pressure, high sensitivity C-reactive protein, LDL cholesterol, HDL cholesterol, total cholesterol,

**Figure 2b.**
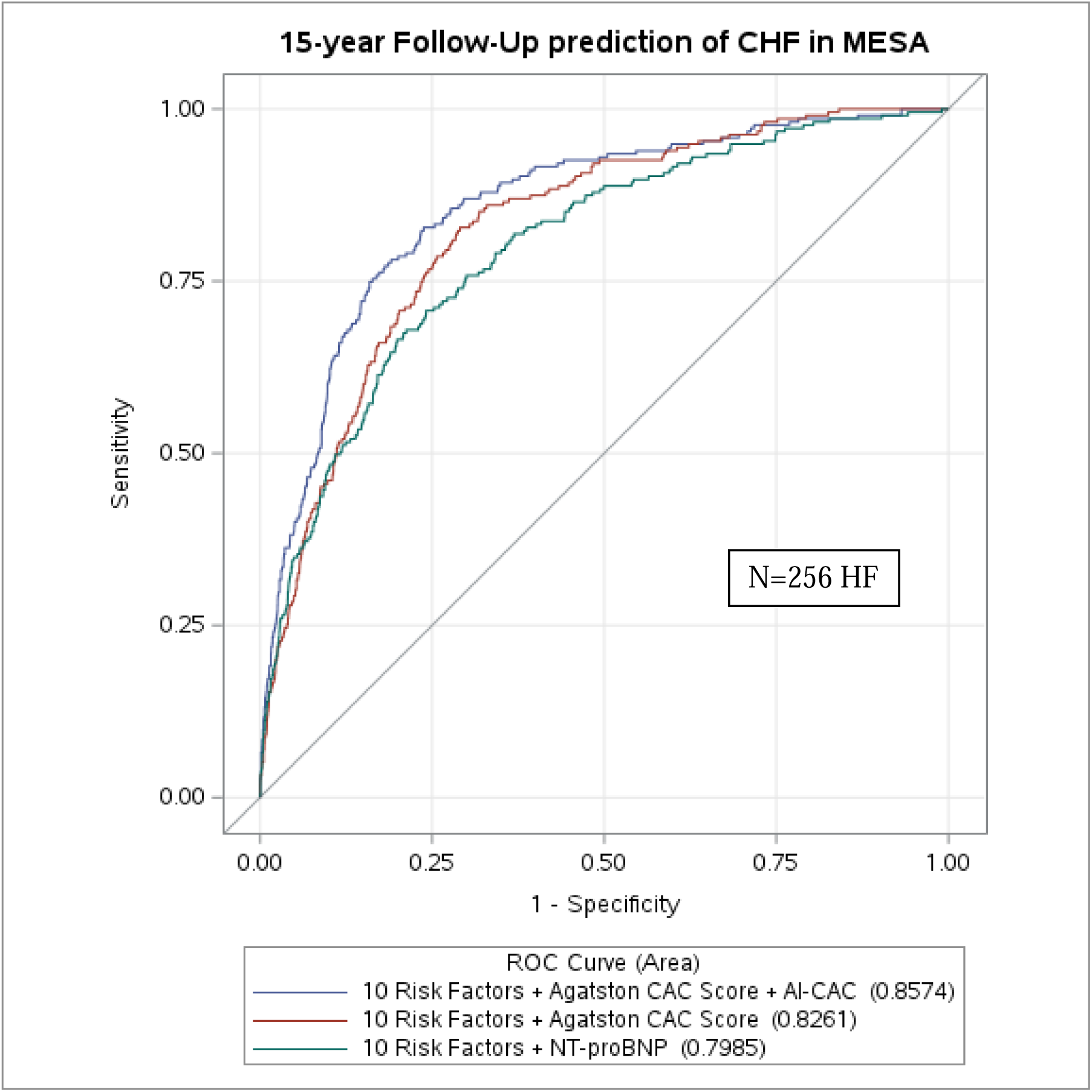
(10 Risk Factors + BNP) vs. (10 Risk Factors + AI-CAC): p=0.0002; (10 Risk Factors + Agatston CAC Score) vs. (10 Risk Factors + AI-CAC + Agatston CAC score): p=0.0005 Risk factors: Age, gender, BSA, diabetes, current smoking, hypertension medication, systolic blood pressure, diastolic blood pressure, high sensitivity C-reactive protein, LDL cholesterol, HDL cholesterol, total cholesterol

**Table 3.** Net Reclassification Index (NRI) of Artificial Intelligence-enabled Cardiac Chambers Volumetry (AI-CAC) added to N-terminal pro-brain natriuretic peptide (NT-proBNP), Agatston CAC score, and Clinical Risk Factors for 5-,10-, and 15-year prediction of heart failure (HF)

| **Adding AI-CAC** | **Net Reclassification Improvement** | **NRI Standard Error** | **Z-Value for NRI** | **NRI P-Value** | **NRI 95% CI** | **% of Events correctly reclassified** | **Event P-Value** | **% of Nonevents correctly reclassified** | **Non-Event P-Value** |
| --- | --- | --- | --- | --- | --- | --- | --- | --- | --- |
| Category-Free NRI for Base Model CAC Score |  |  |  |  |  |  |  |  |  |
| Year 5 | 0.81 | 0.10140 | 6.53119 | <.0001 | (0.488,0.8854) | 26% | 0.0123 | 43% | <.0001 |
| Year 10 | 0.67 | 0.070585 | 9.59314 | <.0001 | (0.5745,0.8512) | 31% | <.0001 | 40% | <.0001 |
| Year 15 | 0.71 | 0.060764 | 10.9591 | <.0001 | (0.5795,0.8177) | 30% | <.0001 | 40% | <.0001 |
| Category-Free NRI for Base Model BNP |  |  |  |  |  |  |  |  |  |
| Year 5 | 0.68 | 0.10195 | 6.29587 | <.0001 | (0.4619,0.8615) | 24% | 0.0218 | 42% | <.0001 |
| Year 10 | 0.71 | 0.070952 | 9.03787 | <.0001 | (0.5312,0.8093) | 29% | <.0001 | 38% | <.0001 |
| Year 15 | 0.68 | 0.060747 | 10.1738 | <.0001 | (0.5255,0.7637) | 28% | <.0001 | 36% | <.0001 |
| Category-Free NRI for<br>Base Model Clinical (10<br>known risk factors)† |  |  |  |  |  |  |  |  |  |
| Year 5 | 0.45 | 0.10307 | 4.57703 | <.0001 | (0.2792,0.6833) | 20% | 0.0606 | 29% | <.0001 |
| Year 10 | 0.50 | 0.073016 | 6.24228 | <.0001 | (0.3207,0.607) | 18% | 0.0131 | 28% | <.0001 |
| Year 15 | 0.43 | 0.06262 | 6.76120 | <.0001 | (0.0378,0.5535) | 17% | 0.0053 | 26% | <.0001 |

† 10 known Clinical risk factors: Age, gender, BSA, diabetes, current smoking, hypertension medication, systolic blood pressure, diastolic blood pressure, high sensitivity C-reactive protein, LDL cholesterol, HDL cholesterol, total cholesterol

A significant number of low-risk categorized participants by CAC score and ASCVD Pooled Cohorts Equation have enlarged cardiac chambers (Figure 3a-b). Examples of three high risk patients with enlarged LA and LV volume with CAC score 0 and CAC below 50^th^ percentile, who are currently categorized as low risk have been provided in Figure 1.

**Figure 3.**
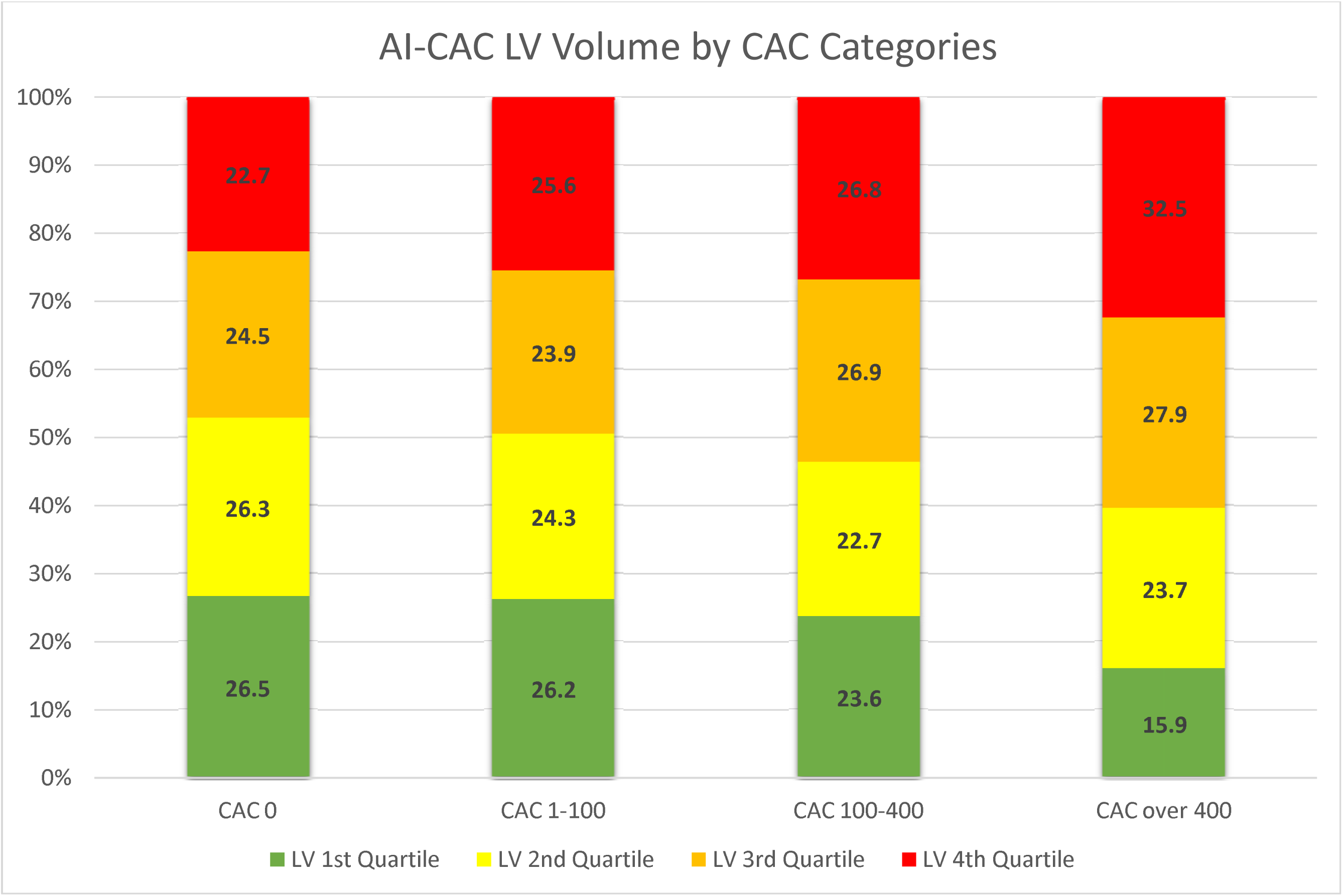
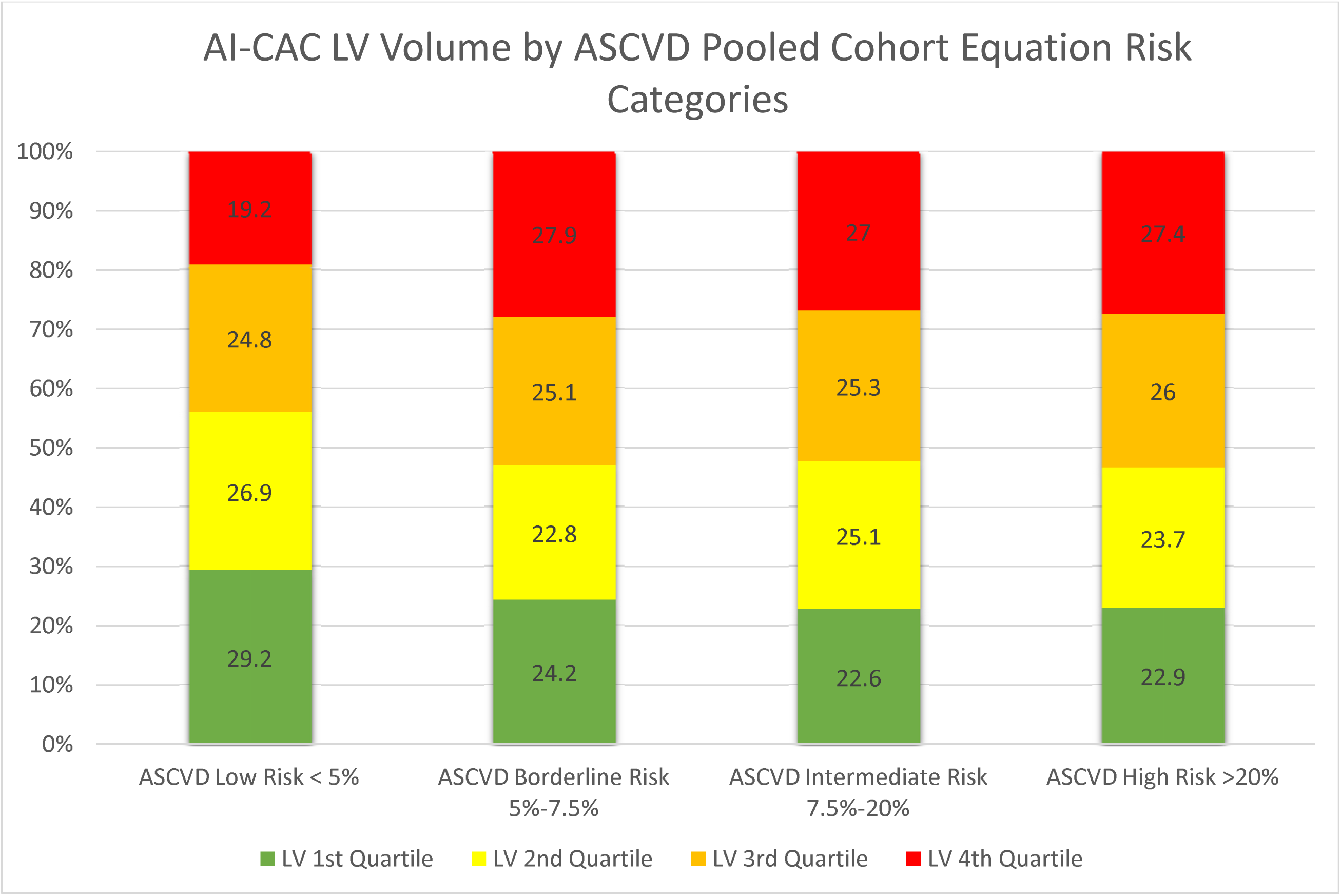
**a-b)** Quartiles of AI-CAC Left Ventricle (LV) Volume by coronary artery calcium (CAC) and ASCVD Pooled Cohorts Equation Categories.

The 125 cases with missing slices were 49.8% male and 50.2% females. None of these cases had a diagnosis of HF, and our investigations did not reveal any association with dependent or independent variables in our study.

## Discussion

To our knowledge this is the first report of an AI-enabled automated cardiac chamber volumetry in non-contrast cardiac CT scans obtained for CAC scoring that predicted future HF in a multi-ethnic asymptomatic population. Our study has shown that the AI-CAC technique non-significantly outperformed NT-proBNP for HF prediction at 5 years, and significantly outperformed BNP at 10 and 15 years. Additionally, it significantly outperformed the Agatston CAC Score for HF prediction at all years and provided a sizable NRI on top of both NT-proBNP and the Agatston Score. We recently reported that the same AI-CAC technique in CAC scans enabled prediction of future AF and outperformed both CHARGE-AF and NT-proBNP for predicting AF at 5, 10, and 15 years^11^. Our studies are not the first to raise awareness on the potential utility of non-coronary findings in CAC scans as they have been reported previously using manual 2D measurements of LV^22,23,24,25^ and LA sizes^26,27,28,29^. However, we provide evidence on the feasibility of using AI for automated 3D volumetry of cardiac chambers that takes on average 20 seconds versus tedious manual measurements that are time consuming and poorly reproducible.

NT-proBNP is a serum biomarker of cardiac volume overload particularly and has been studied extensively in various cardiovascular diseases, in particular heart failure^30,31,32^. Elevated levels of BNP are known to be associated with the presence of heart failure and reduced ejection fraction^33^. Our study shows that AI enabled LV volumetry non-significantly outperformed NT-proBNP for 5-year HF prediction and significantly outperformed NT-proBNP for 10- and 15-year prediction. The small sample size of HF at 5 years may explain p=0.15 at 5yrs despite higher AUC for AI vs NT-proBNP (0.835 vs. 0.795). Additionally, the AI improved NRI of NT-proBNP at all years. It is possible that AI is outperforming NT-proBNP because NT-proBNP is not specific to LV volume overload, whereas AI cardiac chambers volumetry in CAC scans directly measures LV volume.

Recent studies have suggested another opportunistic screening tool using AI-enabled deep learning ECG signals for detection of prevalent ALVD^34,35^. These AI ECG models using a standard 10-second 12-lead ECG data in asymptomatic populations as well as patient populations in clinical datasets have shown to predict future AF^36^ and HF^37^ with comparable accuracy to the HF risk calculators from the ARIC study and the Framingham Heart Study^37^. In another study by Yao et al^38^, ECGs were obtained as part of routine care from a total of 22,641 adults without prior HF. The primary outcome was a new diagnosis of low EF (≤50%) within 90 days of the ECG. The results indicated that use of AI-ECG enabled the early diagnosis of low EF in patients in the setting of routine primary care^38^. We do not have paired data to compare the performance of our AI-enabled cardiac chambers volumetry in CT scans with these AI-enabled deep learning ECG tools for detection of incident AF and HF. However, it is important to note that AI-ECGs are often referred to as “Black-Box AI” meaning the AI output does not show clinicians what features or segments of ECG signals were used for LVH and ALVD detection, whereas in our approach the AI colorfully segments the cardiac chambers in non-contrast CT images where the volumetry measurements are done and enlarged cardiac chambers are detected.

### Non-Coronary findings in CAC Scans

Although there are multiple automated CAC scoring tools available, currently, there is no clinically available tool to clinicians for automated cardiac chambers volumetry in CAC scans. Such measurements are only possible on contrast-enhanced CT scans which require more radiation plus injection of an X-ray contrast agent that is burdensome^39^. Whereas our AI can be applied to any new or existing non-contrast CAC scans for automated cardiac chambers volumetry reports. Other imaging tools including contrast-enhanced cardiac MRI and echocardiography are not suitable as an opportunistic add-on screening tool to CAC scans. Therefore, non-contrast chest CT scans are prime candidates for opportunistic AI-enabled cardiac chambers volumetry for detection of high-risk future AF^11^ and HF. The AI approach can enable automatic screening of over 10 million chest CT scans done each year in the US alone^40^. Such an AI tool can run in the background of radiology PACS and alert providers to cases with enlarged cardiac chambers.

Unfortunately, many high-risk patients with enlarged cardiac chambers, LVH and ALVD are currently undetected, therefore untreated. Early detection of these cases can allow for close monitoring of progression to AF for stroke prevention and intensive medical treatment for HF prevention.

### Limitations

Our study has some limitations. The MESA Exam 1 baseline CT scans, performed between 2000 and 2002, were predominantly conducted using electron-beam computed tomography (EBCT) scanners. This technology is no longer the commonly used method of CAC scanning. Since our AI training was done completely outside of MESA and used a modern multi-detector (256 slice) scanner, we do not anticipate this to affect the generalizability of our findings.

## Conclusion

In this multi-ethnic population study, AI-powered automated cardiac chambers volumetry enabled prediction of HF in CAC scans and improved on the HF predictive value of NT-proBNP and the Agatston Score.

## Data Availability

All data was obtained from MESA Exam 1.

## Clinical Perspectives

The potential value of non-coronary findings in coronary calcium scans is significant. AI-enabled cardiac chambers volumetry can detect patients with enlarged LV and alert providers to take preventive actions. The clinical utility of this opportunistic add-on AI to CAC scans warrants further validation in other longitudinal cohorts.

## Acknowledgement

This research was supported by 2R42AR070713 and R01HL146666 and MESA was supported by contracts 75N92020D00001, HHSN268201500003I, N01-HC-95159, 75N92020D00005, N01-HC-95160, 75N92020D00002, N01-HC-95161, 75N92020D00003, N01-HC-95162, 75N92020D00006, N01-HC-95163, 75N92020D00004, N01-HC-95164, 75N92020D00007, N01-HC-95165, N01-HC-95166, N01-HC-95167, N01-HC-95168 and N01-HC-95169 from the National Heart, Lung, and Blood Institute, and by grants UL1-TR-000040, UL1-TR-001079, and UL1-TR-001420 from the National Center for Advancing Translational Sciences (NCATS). The authors thank the other investigators, the staff, and the participants of the MESA study for their valuable contributions. A full list of participating MESA investigators and institutions can be found at http://www.mesa-nhlbi.org

